# A Protective Haplotype *in trans* Modifies Penetrance and Severity of *TOR1A* Dystonia

**DOI:** 10.64898/2026.09.05.26361794

**Authors:** Roy Dayan, Orli Halstuk, Shira Yanovsky-Dagan, Jonathan Rips, Chaggai Rosenbluh, Ilana Livyatan, Michelle Grunin, Morasha Plesser-Duvdevani, Hagai Bergman, Tamir Ben Hur, James R Lupski, Deborah Raymond, Susan Bressman, Rachel Saunders-Pullman, Aloysius Domingo, Laurie J Ozelius, Shai Carmi, Tamar Harel, David Arkadir

## Abstract

**Introduction:** Incomplete penetrance and variable expressivity characterize autosomal dominant neurological disorders, but their genetic determinants remain largely unknown. DYT-*TOR1A* dystonia provides a model for investigating incomplete penetrance. We aimed to identify modifiers of disease penetrance and severity beyond the known *TOR1A* c.646G>C modifier.

**Methods:** In a prospective genotype–phenotype association study, we phenotyped Ashkenazi Jewish carriers of *TOR1A* c.907_909del using a prespecified five-level severity score. The primary analysis compared participants with no or minimal dystonia (scores 0–1) and severe dystonia (scores 3–4), after excluding c.646G>C carriers. Findings were assessed in an independent retrospective cohort. Sanger, exome, and short-read and long-read whole-genome sequencing were used for genotyping.

**Results:** The prospective cohort included 61 carriers: 21 had severe dystonia and 28 had no or minimal dystonia, six of whom carried c.646G>C. Among the 43 participants in the primary analysis, haplotype A was identified in 33 (77%) and haplotype B in eight (19%). All haplotype B carriers had no or minimal dystonia (p=0.003). Haplotype B remained associated with a milder phenotype after including 12 participants with intermediate severity (p=0.006). The retrospective cohort included 147 carriers, 16 of whom carried c.646G>C. After their exclusion, dystonia manifested in 12 (38%) of 32 participants with haplotype B and 63 (64%) of 99 without it (p=0.009). Haplotype B was also associated with lower severity scores (p=0.011). Together, c.646G>C and local haplotype status predicted the phenotype of more than two-thirds of carriers. In silico analyses linked haplotype B to lower *TOR1A* and higher *TOR1B* expression in dystonia-relevant brain regions.

**Conclusion:** A common local haplotype *in trans* to the disease-causing *TOR1A* variant was strongly associated with reduced penetrance and severity. These findings establish DYT-*TOR1A* dystonia as a model for investigating how local genetic variation contributes to incomplete penetrance in autosomal dominant neurological disorders.

## INTRODUCTION

The underlying mechanisms of incomplete penetrance and phenotypic severity (variable expressivity) in genetic disorders remain poorly understood.^1–3^ Earlier research emphasized the influence of environmental factors on penetrance and disease severity.^4^ Emerging evidence, however, highlights the role of genetic modifiers, either through the cumulative effect of multiple weak genetic modifiers^5^ or through a single strong modifier.^5–9^ Such a strong modifier can include single-nucleotide variants, as well as structural variations.^8,10,11^ Although incomplete penetrance is common in autosomal dominant diseases, the genetic modifiers underlying this variability remain largely unidentified.^1,3^

Attempts to identify genetic modifiers of incomplete penetrance and severity face several challenges.2,3 Different pathogenic variants in the culprit gene are associated with variable penetrance and disease severity, complicating efforts to isolate modifiers.12 Studied populations, particularly in rare diseases, remain small relative to the extensive number of candidate variants within the human genome.13–15 In addition, age-dependent penetrance impedes modifier detection, as it is often unclear whether a non-manifesting individual would develop the disease later in life.16 For these reasons, dystonia caused by a pathogenic *TOR1A* deletion (DYT-*TOR1A*, formerly DYT1) represents an ideal model for unraveling genetic modifiers of penetrance.

DYT-*TOR1A*, a dominantly inherited dystonia with approximately 30% penetrance,^17^ exhibits several unique characteristics. Unlike most genetic disorders, nearly all cases are caused by the same three-nucleotide deletion, NM_000113.3:c.907_909delGAG, which removes one of two consecutive glutamate residues (p.Glu303del).^17,18^ This deletion is markedly more frequent among Ashkenazi Jews, a population shaped by a historical bottleneck that resulted in a limited number of haplotypes.^19^ Moreover, the disease rarely manifests after the age of 26 years,^20^ allowing a relatively clear distinction between manifesting and non-manifesting carriers. These characteristics make DYT-*TOR1A* a useful model for investigating genetic modifiers of penetrance and severity.

A strong genetic modifier for DYT-*TOR1A* was previously identified within the *TOR1A* gene, located in *trans* (a term that will be used here to indicate only the opposite homologous allele and not a distant element) to the causative deletion (c.646G>C, p.Asp216His).8 However, because this variant occurs in only 20% of carriers,21 a substantial proportion of the variable penetrance remains unexplained.

To investigate whether additional genetic elements contribute to incomplete penetrance in DYT-*TOR1A*, we studied two cohorts of well-characterized manifesting and non-manifesting carriers of c.907_909del in *TOR1A*. Our analysis revealed a haplotype surrounding *TOR1A*, *in trans* to the pathogenic deletion, that confers strong protection against dystonia, suggesting that local genetic variation accounts for the observed phenotype in the majority of DYT-*TOR1A* cases.

## METHODS

### Study participants

We conducted an observational genotype-phenotype association study of carriers of the *TOR1A* c.907_909del pathogenic variant and their relatives recruited at the Movement Disorders clinic, Hadassah Medical Center (Jerusalem, Israel) between 2016 and 2025. See supplementary methods for detailed methods. Inclusion criteria were (1) clinically manifest dystonia in individuals aged 16 years or older or (2) absence of dystonia in individuals older than 26 years, since onset after that age is uncommon in DYT-*TOR1A*.^20,22^ All participants provided written informed consent. The study was approved by the Hadassah Medical Center Institutional Review Board and the Israeli Ministry of Health supreme ethics committees (HMO-0096-15).

### Clinical evaluation

Participants were examined by a movement disorders specialist. Because many severely affected participants had already improved after deep brain stimulation^23,24^ and examination at recruitment was therefore not expected to reflect lifetime disease burden, we reviewed pre-DBS records and incorporated these into a prespecified 5-level maximum severity score reflecting the natural history of DYT-*TOR1A* (Table S1) rather than using a conventional scale at the time of recruitment (e.g., Fahn-Marsden dystonia rating scale^25^). In our scale, scores 0-1 represented none or minimal dystonia, score 2 intermediate dystonia without lower-limb involvement, and scores 3-4 generalized dystonia with impaired or lost independent ambulation, respectively. The prespecified primary analysis compared participants at phenotypic extremes (scores 0-1 vs. 3-4) after exclusion of *TOR1A* c.646G>C carriers.

### Genetic and Secondary Analyses

All participants underwent Sanger sequencing to confirm *TOR1A* c.907_909del. Carriers were additionally genotyped for *TOR1A* c.646G>C. Selected participants underwent whole-exome sequencing, short-read whole-genome sequencing (WES and WGS), and Oxford Nanopore Technologies (ONT) long-read whole-genome sequencing (Figure S1) to identify and phase variants around the *TOR1A*/*TOR1B* locus and define haplotypes in trans to the pathogenic deletion. As a secondary analysis, polygenic risk scores for Parkinson disease^26^ and laryngeal dystonia^27^ were calculated in a subset of c.646G>C-negative participants genotyped on single-nucleotide polymorphism arrays after imputation.

### Independent Replication Cohort

The cohort included previously reported *TOR1A* c.907_909del carriers with clinical assessment and genotyping methods also described previously.^8,28^ Data were harmonized with the current clinical scoring independent of the genetic data. Non-Jewish *TOR1A* c.907_909del carriers were excluded from this analysis because haplotypes could not be clearly inferred in this group (Table S3).

### Functional Annotation and Supportive Expression Studies

Population frequency, linkage disequilibrium, and estimated evolutionary age of haplotype-defining variants were assessed using public genomic databases.^21,29–31^ Predicted tissue-specific expression effects were examined using expression quantitative trait locus data from Genotype-Tissue Expression v8 (GTEx)^32^ and AlphaGenome^33^ for sequence-based regulatory prediction from long-read haplotype sequences. Supportive laboratory studies assessed *TOR1A* and *TOR1B* expression in peripheral blood and in patient-derived lymphoblastoid cell lines representing the main haplotype groups. Promoter-region methylation profiles were extracted from ONT data.

### Statistical analysis

The primary outcome was dystonia severity. The primary analysis compared haplotypes *in trans* between participants with minimal/no dystonia (scores 0-1) and severe dystonia (scores 3-4) after excluding carriers of c.646G>C. Secondary analyses incorporated the full severity scale across all carriers and evaluated the combined effects of the protective haplotype and c.646G>C. Categorical variables were compared using χ² or Fisher exact tests, and continuous variables using Student’s t tests or Mann-Whitney U tests, as appropriate; Kruskal-Wallis testing was used for comparisons across more than 2 groups. For regression analyses, symptom severity beyond minimal dystonia (score >1) was modeled as the dependent variable, with protective variation *in trans* and sex as independent variables; mixed-effects logistic regression with family as a random effect was used as a sensitivity analysis to account for pedigree structure. Because the primary analysis was prespecified and focused on local genetic variation near *TOR1A*, no genome-wide multiple-testing framework was applied; secondary annotation and functional analyses were considered supportive or exploratory. All tests were 2-sided with α=.05. Statistical analyses were performed in R2024.12.1 and PLINK2.0.^34^ Additional analytical details are provided in the supplementary methods.

## RESULTS

### Participant Recruitment and Characteristics

We recruited 73 individuals from 21 unrelated families in which at least a single individual had symptomatic DYT-*TOR1A* secondary to c.907_909del. Sanger sequencing confirmed the c.907_909del variant in 61/73 participants (mean age at consent 30.9 years, range 16-66 years, 36.0% females).

Phenotypic assessment of carriers revealed that 28/61 (45.9%) exhibited no or only minimal dystonia (minDys; severity score 0–1, see Methods), whereas 21/61 (34.4%) were classified as having severe dystonia (svrDys; severity score 3–4). These two groups, representing the extremes of the DYT-*TOR1A* phenotypic spectrum, were used to stratify individuals in discovery analyses. An intermediate group of 12/61 (19.7%) individuals with moderate dystonia (intDys, severity score 2) was excluded from the primary analysis according to the predefined study design.

The severity score mirrored the expected disease course of DYT-*TOR1A*: higher scores were associated with earlier onset (p=0.032), greater use of neurosurgical procedures (p<0.001), and lower frequency of the known protective variant c.646G>C (p=0.024, Table 1). The presence of the known c.646G>C modifier explained the phenotype in 6/28 individuals in the minDys group. To avoid masking a novel modifier associated with minimal phenotype, we excluded these six individuals from subsequent analyses based on our a priori study design.

**Table 1:** Participant characteristics of the prospective discovery cohort.

| Severity score | minDys (score 0-1, n=28) | intDys (score 2, n=12) | svrDys (score 3-4, n=21) | P-value (3-way) | P-value (minDys vs. svrDys) |
| --- | --- | --- | --- | --- | --- |
| Sex (F/M) | 15/13 | 6/6 | 5/16 | 0.096 | **0.036** |
| DBS/Pallidotomy | 0 (0%) | 2 (16.6%) | 18 (85.7%) | **<0.001** | **<0.001** |
| Mean age of onset in years ±SD (range) | Score 0: N/A<br>Score 1: 19 ± 1.4 (18-20, N = 2)* | 25 ± 23.4 (7-60, N = 11) | 10.2 ± 3.2 (5-19, N = 21) | **0.039** | **0.032** |
| c.646G>C | 6 (21.4%) | 1 (8.3%) | 0 (0%) | 0.062 | **0.024** |

Abbreviations: DBS: Deep Brain Stimulation. SD: Standard deviation. minDys: minimal dystonia. intDys: intermediate dystonia. svrDys: severe dystonia. N/A: not available. \* Data regarding age of onset among the minDys group was only partially available since most patients were unaware of mild signs or could not recall a specific onset. Significant p-values are in bold.

### Distant Genetic Factors and Proximal *cis* Variation Do Not Explain Incomplete Penetrance of c.907_909del

Screening of dystonia-related genes (Table S4) in WES data from c.646G>C-negative participants with minDys (n=22) and svrDys (n=21) identified no pathogenic or likely pathogenic variants (see methods) explaining penetrance differences. We also searched for pathogenic variants in genes associated with parkinsonism because of the overlap between these phenotypes,^35^ identifying *LRRK2* G2019S in two participants with svrDys. Because neither had parkinsonian signs, the significance of this finding remains uncertain.

We next tested whether polygenic burden can differentiate minDys and svrDys carriers. As no PRS was available for generalized dystonia, we tested published PRS for laryngeal dystonia^27^ and Parkinson disease^26^ in c.646G>C-negative participants. Both analyses were negative (Figure S2A-B; p=0.332 and p=0.491, respectively).

Marked vertical and horizontal phenotypic variability within families argued against a *cis*-acting modifier linked to c.907_909del. Still, we interrogated a 50-kb cis region spanning *TOR1A*, *TOR1B*, and nearby regulatory elements. WES (n=43) and long-read sequencing (n=15) identified a single shared haplotype with only sporadic variants that did not distinguish severity groups (Figure S3), consistent with prior studies.^36^

### A distinct haplotype in *trans* of *TOR1A* c.907_909del is associated with reduced penetrance and milder phenotype of DYT-*TOR1A*

Local genetic variations often modify penetrance and phenotypes.^37^ For this reason, we next searched for modifiers within the region of *TOR1A*, other than the known *TOR1A* c.646G>C modifier. To increase statistical power and reduce false negatives due to multiple-comparison correction, we analyzed haplotypes (linked variants) rather than individual variants.

This analysis identified two main haplotypes in *trans* to *TOR1A* c.907_909del in our cohort (Figure 1A, excluding carriers of *TOR1A* c.646G>C). First, the major haplotype (haplotype A) was found in 33/43 (76.7%) individuals from the minDys and svrDys groups. Second, a distinct minor haplotype (haplotype B), defined by multiple linked variants (Table S5), was observed in 8/43 individuals (18.6%). Haplotype B spanned a region of approximately 45-54 kb encompassing *TOR1B*, *TOR1A*, *C9orf78* and part of *USP20* (chr9: 129790096-129834742 in hg38). In addition to these two haplotypes, the *trans* haplotype in two individuals (2/43, 4.7%) was hybrid, incorporating variants from both haplotypes (mixed haplotype, Table S9). Haplotype distinction was further confirmed using principal component analysis with unsupervised clustering (Figure 1B). The c.646G>C variant completely clustered with haplotype A (Figure S4).

**Figure 1:**
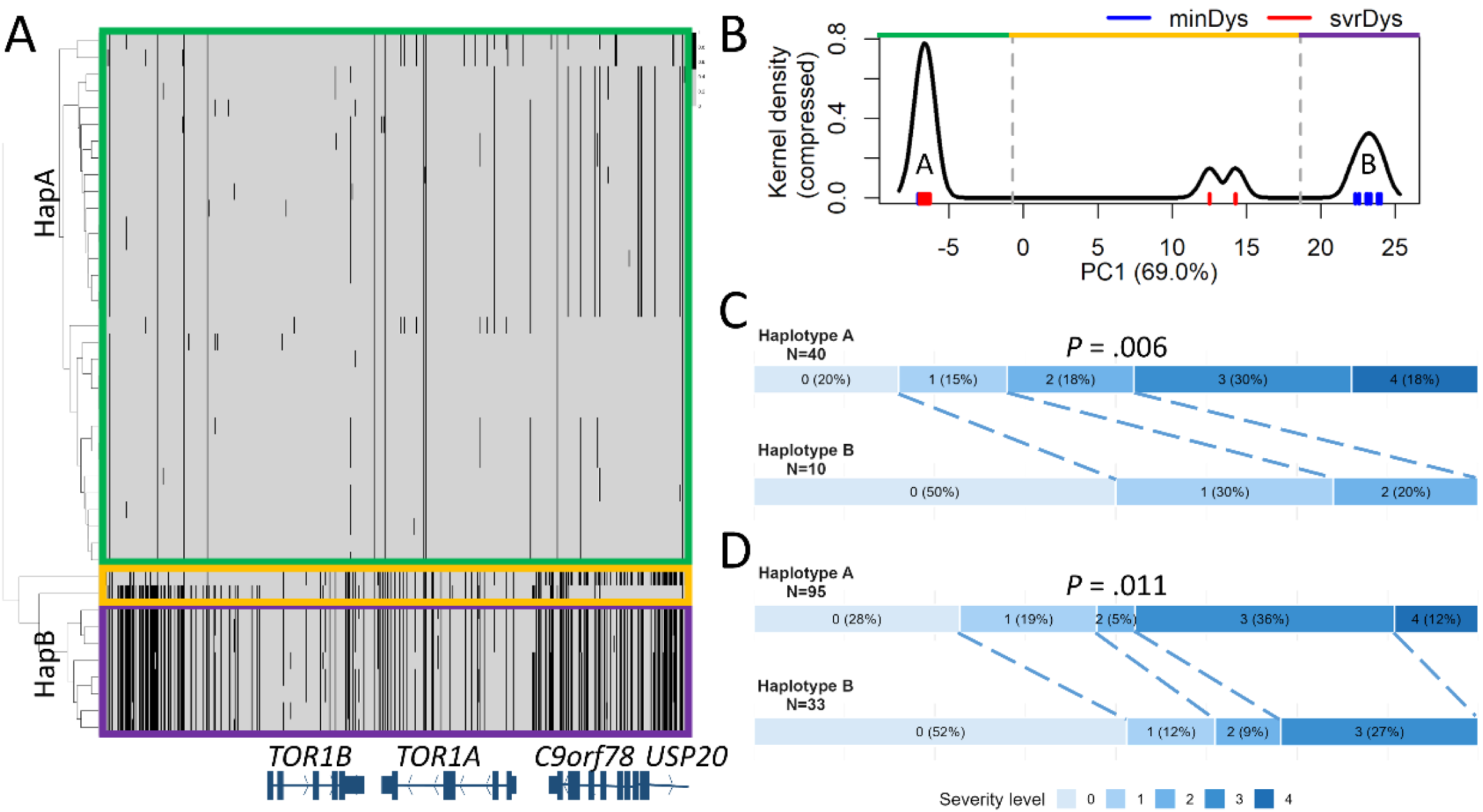
A common minor haplotype *in trans* to *TOR1A* c.907_909del is associated with reduced penetrance of DYT-*TOR1A*. A) Heatmap of all minDys and svrDys participants without c.646G>C variant using unsupervised clustering shows two distinct haplotypes: Haplotype A (green box), the major one in our cohort, and a minor Haplotype B (purple). In addition, a mixed haplotype was observed in two individuals (yellow). The heatmap shows variants only (unfiltered), so their positions within the 50-kb window are not to scale. Grey indicates the reference allele; black rectangles indicate the non-reference allele. B) Kernel density plot of the first principal component (covering 69.0% of variance) showing the separation of these haplotypes. C) A comparison of dystonia severity levels between the two main haplotypes shows lower severity levels among participants with Haplotype B (median 0.5, n=10) than among patients with Haplotype A (median 2, n = 40, p=0.006). Individuals with a mixed haplotype (n=4) or with c.646G>C (n=7) are excluded from this analysis. D) A comparison of dystonia levels in a retrospective independent cohort of Ashkenazi Jewish *TOR1A* c.907_909del carriers shows lower severity levels among participants carrying three variants from Haplotype B (median 0, n=33) than among participants not carrying these variants (median 2, n=95, p=0.011). Participants with c.646G>C were excluded from this analysis. Notably, because this cohort was genotyped by Sanger sequencing for specific variants only, and because the same variants were also observed in individuals with a mixed haplotype in the prospective cohort, some participants classified as Haplotype B carriers in the retrospective cohort may instead carry a mixed haplotype. Finally, in a combined analysis of both Ashkenazi Jewish cohorts excluding carriers of the known protective *TOR1A* c.646G>C variant, Haplotype B remained strongly associated with lower dystonia severity, even when mixed-haplotype carriers were conservatively classified as Haplotype B carriers (n=203; p=0.003). Abbreviations: HapA: Haplotype A (major). HapB: Haplotype B (minor). minDys: minimal dystonia. svrDys: severe dystonia. PC1: the first principal component.

Examining dystonia severity among individuals with each of the two main haplotypes revealed that Haplotype B is associated with a strong protective effect. None of the 8 individuals carrying Haplotype B were in the svrDys group, whereas 19/33 (57.6%) of patients carrying Haplotype A were in this group (p=0.003). Notably, the association with reduced dystonia remained significant even when individuals carrying the mixed haplotype, were classified as having Haplotype B (p=0.037).

To examine the protective effect of Haplotype B on all severity scores, we analyzed the group as a whole (n=61), including patients with intermediate levels of symptoms (intDys, n=12). Analyzing the level of dystonia as a continuous variable also revealed a strong protective effect of Haplotype B (median 0.5) compared to Haplotype A (median 2, p=0.006, Figure 1C). Overall, phenotype status was concordant with the presence or absence of a protective factor *in tran*s (either c.646G>C or haplotype B) in 44/61 carriers (72.1%).

### Variants from Haplotype B are associated with reduced penetrance and milder disease in an independent replication cohort

To replicate our findings, we retrospectively analyzed an independent cohort of Ashkenazi Jewish *TOR1A* c.907_909del carriers (n = 147) who had been genotyped for three of the variants within Haplotype B (Table S3). These variants were in complete linkage disequilibrium and they were either all present or all absent in this cohort.

After excluding 16 individuals with the known protective c.646G>C variant, clinical dystonia was more often reported in individuals without Haplotype B compared to individuals with Haplotype B (63/99 vs. 12/32 respectively, p=0.009). These results are concordant with the presence or absence of a protective factor *in trans* (c.646G>C or Haplotype B–associated variants) in 98/147 (66.7%) individuals.

Rating of dystonia at maximum distribution for the replication cohort were collapsed into the categories of the severity-scoring system (n=128 participants with available data). Analysis of dystonia severity as a continuous variable replicated our results by demonstrating a protective effect of Haplotype B (p=0.011; Figure 1D).

In a logistic regression analysis of the combined Ashkenazi Jewish cohorts with clinically-significant dystonia (severity score ≥2) as the dependent variable, the two mutually exclusive protective in trans elements, c.646G>C and Haplotype B,8 were each independently associated with lower severity (p=0.001 and p=0.030, respectively).

Male sex showed a signal toward higher severity, but this did not reach significance (OR, 1.67; 95% CI, 0.93-3.00; p=0.094; Table S7). Results were unchanged in mixed-effects models accounting for family structure.

### Origin of Haplotype B

Haplotype B comprised two linked sub-haplotypes (Figure S5, Table S5). Variants in an older *TOR1A*-centered component were dated to approximately 1.1 million years ago and are present across modern human populations and in sequenced Neanderthals and Denisovans, indicating origin before the modern-archaic split. These variants are most frequent in Ashkenazi Jews (mean allele frequency 27.9%) but are also common in Europeans (24.4%), Africans (22.0%), and East Asians (18.3%).^38^ A more recent *TOR1B*-centered component included variants dated to approximately 269,000 years ago and occurring at similar frequencies in Ashkenazi Jews and Europeans (19.7% and 20.5%), slightly less in East Asians (16.4%), and much less in Africans (4.7%), suggesting fusion of the two sub-haplotypes in the ancestors of present-day non-Africans.

### Haplotype B is associated with downregulation of *TOR1A* and upregulation of *TOR1B* in the brain in silico

Because *TOR1B* overexpression protects against DYT-*TOR1A* in animal models,^39^ we assessed the predicted effect of haplotype B on *TOR1A* and *TOR1B* expression. In GTEx, haplotype B variants were associated with lower *TOR1A* and higher *TOR1B* expression across multiple brain regions, with the strongest effects in cerebellum and striatum (adjusted p<0.001; Figure 2A-B, Tables S5 and S8). Across the 200-kb region surrounding c.907_909del, these variants mapped to a single LD-block accounting for most of the brain eQTL signal (Figure S6). In the two participants with mixed haplotypes, several key tissue-relevant eQTLs associated with reduced *TOR1A* and increased *TOR1B* expression were absent (Table S9). In AlphaGenome analyses of 1Mb long-read sequences from 12 participants (2 haplotype B, 10 haplotype A), haplotype B was likewise associated with a lower predicted *TOR1A/TOR1B* expression ratio in cerebellum, caudate, and putamen (all p<0.001; Figure 2C). Although fresh brain tissue from individuals with DYT-*TOR1A* was unavailable and iPSC-derived neuronal models were not generated. We explored the effects on expression using available patient-derived peripheral blood and lymphoblastoid cell lines, and observed suggestive evidence of Haplotype B-associated modulation of the mutant *TOR1A* to *TOR1B* expression ratio (Figure S7).

**Figure 2:**
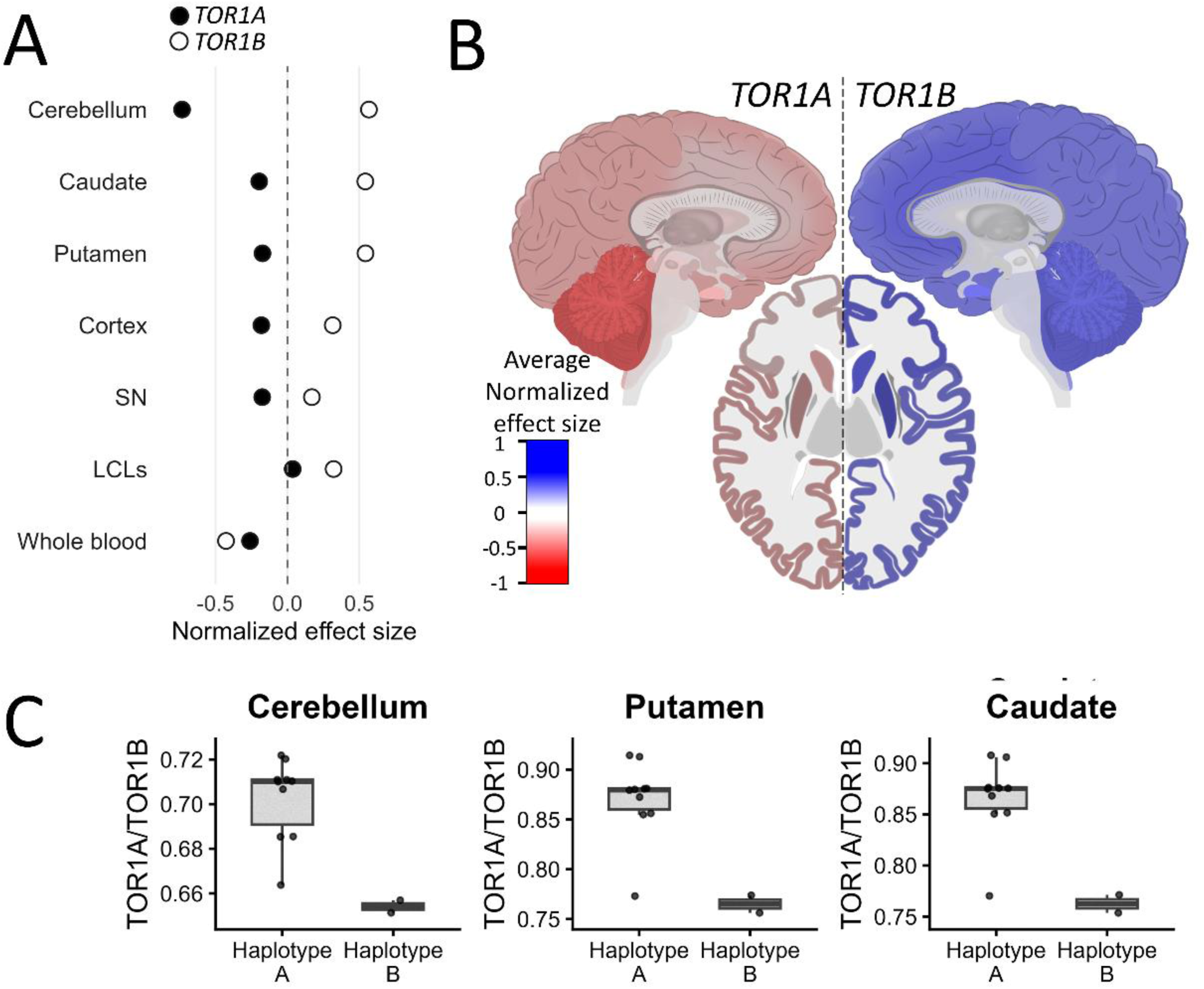
Haplotype B is associated with changes in *TOR1A* and *TOR1B* expression in relevant tissues. A–B) Average normalized effect size (NES) extracted from the GTEx expression Quantitative Trait Loci (eQTL) calculator across brain regions and cell lines shows broad downregulation of *TOR1A* and upregulation of *TOR1B*, most pronounced in dystonia-related regions (cerebellum and basal ganglia). C) AlphaGenome-based *in silico* prediction using blood-derived 1-Mb long-read sequences *in trans* to *TOR1A* c.907_909del showed a significantly lower predicted *TOR1A*/*TOR1B* expression ratio in cerebellum, caudate, and putamen among Haplotype B carriers (n=2) compared with Haplotype A carriers (n=10; p<0.001). The central black line in the boxplot marks the median value; the box spans the interquartile range (IQR; 25th–75th percentiles).

## DISCUSSION

We prospectively studied here 61 individuals with the *TOR1A* c.907_909del pathogenic variant, who were either non-manifesting carriers or patients with clinical manifestations of DYT-*TOR1A*. For this aim we used a clinical scale reflecting the natural history of the disease. Genetic analysis revealed a unique 45-54 kb haplotype in *trans* to c.907_909del (Haplotype B) that is strongly associated with a protective effect from severe dystonia (p=0.003). An independent retrospective cohort, of 147 carriers, replicated these findings. In both cohorts, this protective haplotype, together with the known *TOR1A* c.646G>C protective variant *in trans*, seem to account for two-thirds of the observed penetrance among DYT-*TOR1A* c.907_909del carriers. The concordant findings in two independent large cohorts support the robustness of this association and its potential utility for genetic counseling in the appropriate clinical setting.

Identifying genetic modifiers in monogenic diseases is constrained by uncertainty regarding whether non-manifesting carriers or carriers without subjective symptoms (but with minimal signs on examination) will eventually develop severe disease.16 The lifelong trajectory of most individuals with *TOR1A* c.907_909del is largely predictable - those who do not manifest dystonia by the age of 26 will remain mostly asymptomatic, while those who develop early-onset dystonia involving the lower limbs will develop generalized, disabling dystonia.20 A small subgroup of individuals with dystonia not involving the lower limbs has a less predictable trajectory.22 To improve our ability to detect genetic modifiers in a relatively small group of patients and to overcome the strong symptomatic effect of DBS, we used a clinical scale correlated with the lifelong disease trajectory, rather than the classical scale that documents signs and symptoms at a specific point in time.

Although the characteristics of DYT-*TOR1A* make it an ideal model for studying penetrance and variable expressivity, it remains a rare disease. As such, the number of carriers available for study is disproportionately small relative to the number of candidate variants in the human genome.^14^ To address this challenge, we focused on the genomic vicinity of the pathogenic locus. The genomic vicinity of a culprit gene has been shown to harbor modifiers in DYT-*TOR1A*^8^ and in Spinal Muscular Atrophy (SMA).^40^ This challenge was also addressed by analyzing the overall haplotype rather than individual variants.^41^ Our approach enabled us to identify a minor haplotype (Haplotype B) that functions as a strong protective modifier against severe dystonia. The large number of variants in Haplotype B, which are in linkage disequilibrium, makes it difficult to pinpoint which variant, or set of variants, underlies the protective effect. This pattern of *trans*-modifying haplotypes has likewise been reported in other diseases.^42–44^ Interestingly, two patients with different mixtures of haplotype A and B developed severe dystonia, suggesting that the protective effect is mediated by the cumulative effect of several variants.^41,45^

*In silico* results suggest that Haplotype B is associated with lower expression of *TOR1A* combined with higher expression of its paralog, *TOR1B*, in dystonia-relevant brain tissues. TorsinB, encoded by *TOR1B*, shares 70% sequence identity with torsinA and shows reciprocal developmental expression,^18,46^ suggesting a candidate mechanism for its protective effect. Indeed, in a DYT-*TOR1A* mouse model, *TOR1B* knockout exacerbates dystonia, whereas overexpression prevents manifestation.^39^ This paralog-based compensation mirrors mechanisms described in other diseases, including SMA, in which *SMN2* copy number modulates the severity of *SMN1*-related disease,^40^ and Duchenne muscular dystrophy, in which utrophin upregulation can partly compensate for dystrophin deficiency.^47^ While our *in vitro* results support the *in silico* ones, future validation in induced disease-relevant neuronal cells would serve as additional strong support. Still, as current preclinical therapeutic strategies in DYT-*TOR1A* focus on allele-specific inhibition of the mutant *TOR1A* allele,^48^ our findings indicate that compensation by upregulation of *TOR1B*^39^ may provide a complementary approach.

Incomplete penetrance and variable expressivity were traditionally attributed primarily to environmental factors.^4,49^ Cumulative evidence, however, indicates that genetic determinants play an increasingly central role in the understanding of these phenomena.^5,6,49^ This study highlights how bioinformatics tools tailored for the optimal disease model can reshape our understanding of penetrance and disease severity. As tools for interrogating noncoding variation and long-range regulatory interactions continue to improve,^33^ prediction of the clinical consequences of *TOR1A* c.907_909del may become increasingly precise, further shrinking the unexplained component of penetrance currently attributed to environmental factors.^50^

## AUTHOR CONTRIBUTIONS

RD, LJO, HB, JRL, SC, TH and DA were involved in the study design. RD, OH, SYD, JR, CR, IL, MPD, JRL, DR, SB, RSP, AD, LJO, TH and DA were involved in data acquisition. RD, MG and SC were involved in statistical analysis. All authors were involved in data interpretations. RD and DA wrote the first draft of the manuscript. All authors critically reviewed drafts of the manuscript, approved the final version for submission, and vouch for all aspects of the work. All authors had full access to all participant-level study data listings in addition to generated analyses and outputs. All authors accept responsibility to submit for publication.

## DATA SHARING

Deidentified participant-level data will be made available on reasonable request to the corresponding author, subject to ethics and privacy protections. Because the data include sensitive genetic information, they will not be publicly posted.

## DECLARATION OF INTERESTS

TH and DA were supported by the Dystonia Medical Research Foundation (DMRF). RD was supported by the Israeli Center for Addictions and Mental Health (ICAMH). RD and DA were supported by the Berel and Agnes Ginges Family Foundation. JRL was supported by the United States National Institutes of Health UO1 HG001798 and R35 NS105078.

## ACKNOWLEDGEMENTS

We dedicate this work to the memory of the late Mrs. Frances and Mr. Samuel Belzberg, co-founders of the Dystonia Medical Research Foundation, who devoted their lives to advancing our understanding of dystonia and developing effective treatments.

## Notes

### Competing Interest Statement

The authors have declared no competing interest.

### Author Declarations

The study was approved by the Hadassah Medical Center Institutional Review Board and the Israeli Ministry of Health supreme ethics committees (HMO-0096-15)

