## Supplement for "A Protective Haplotype *in trans* Modifies Penetrance and Severity of *TOR1A* Dystonia"

### SUPPLEMENTARY METHODS

#### Clinical evaluation

All participants were clinically examined by a movement disorders specialist. While designing this study, we were aware that at the time of clinical examination many of the participants with severe dystonia had already improved dramatically with deep brain stimulation (DBS) and therefore using a conventional severity scale, e.g. the Fahn-Marsden dystonia rating scale,[1] at the time of recruitment was not expected to reflect the true severity of their disease. In addition, the severity of dystonia could also not be reliably assessed by briefly turning off the stimulation for ethical reasons because of either a long-term stimulation effect or immediate dystonia rebound.[2,3] To address this limitation, and based on the known lifelong natural history of DYT-*TOR1A*, we developed a novel severity score combining clinical history, including pre-DBS disease severity, with findings from the current examination, see appendix table S1.

#### Sanger sequencing

All study participants underwent Sanger sequencing for the c.907_909del variant, and those who were positive were additionally genotyped for the c.646G>C modifier in *TOR1A*.[4] Additional sequencing was performed for variants that define the haplotype in case of low coverage (appendix table S2). DNA samples, obtained from peripheral blood, were amplified using Dideoxy chain termination method, BigDye Terminator kit v.1.1 cycle, (Applied Biosystems, United States). The amplified products were filtered by Gel filtration cartridges (Edge bio, United States) and analysed on the ABI 3130xl Genetic Analyzer (Applied Biosystems, United States). Primer sequences can be found in appendix table S2.

#### Exome sequencing (ES)

DNA samples, obtained from peripheral blood, were prepared and processed for ES. Briefly, exonic sequences were enriched in the DNA sample using xGen Exome Hyb Panel v2 (Integrated DNA Technologies, Coralville, IA, USA) and sequenced at Hadassah Medical Center using a NovaSeq6000 sequencing system (Illumina, San

Diego, CA, USA). Samples sequenced at the Human Genome Sequencing Center (HGSC) at Baylor College of Medicine underwent exome capture with the BCM-HGSC VCRome 2.1 design (24Mb, Roche NimbleGen), followed by sequencing on the Illumina HiSeq 2000 platform (Illumina, San Diego, CA, USA). Reads were aligned to the reference human genome assembly hg19 (GRCh37) using the Burrows–Wheeler Aligner (BWA) v 0.7.10.[5] Variants were called using the GATK-lite pipeline v48[6] with validation using DNAnexus (Palo Alto) using default parameters with the human genome assembly hg19 (GRCh37) as reference. The FASTQs were uploaded into the Geneyx (previously TGex) Analysis platform[7] with targeted genetic testing (appendix table S4). Genomic coordinates for variants of interest were converted from the hg19 to the hg38 reference assembly.

Whole genome sequencing (WGS)

Genomic DNA was extracted from peripheral blood and Next Generation Sequencing (NGS) libraries were prepared with an Illumina PCR-free TruSeq DNA Library Prep Kit. Sequences were generated on an Illumina NovaSeq 6000 sequencing platform as 161 bp paired-end reads, to a final depth of 30X coverage. The FASTQs were uploaded into the Geneyx Analysis platform.[7] Alignment and variant calling of single nucleotide variations, structural variants, CNVs and repeats were called using Illumina DRAGEN Bio-IT. The resulting VCF files were comprehensively annotated on the Geneyx Analysis annotation engine, and presented for analysis, filtering and interpretation.

Long-read WGS

Long-read WGS (Oxford Nanopore technologies, ONT) was performed as previously described.[8] Briefly, genomic DNA was extracted from peripheral blood (Qiagen Blood and Cell Culture DNA Kit), quantified by NanoDrop and Qubit dsDNA HS, mechanically sheared (FastPrep-96), normalised to 2-3 µg in 50 µl, and libraries were prepared with SQK-LSK114 and the NEBNext Companion Module (E7180S) according to the manufacturer's protocols. Libraries were loaded onto R10.4 flow cells and sequenced on a PromethION48 for 72 h with high-accuracy basecalling and 5-mC methylation calling enabled. Reads were basecalled with Guppy (v6.3.9), quality-controlled and aligned to GRCh38 with minimap2;[9] variants were called with Clair3 (SNVs/indels) and Sniffles2 (SVs).[9,10]

Polygenic risk score (PRS)

Blood samples from 39 participants (20 minDys, 19 svrDys) who were negative for the known c.646G>C modifier were genotyped using Axiom Precision Medicine Diversity Research Array (Applied Biosystems) with >850,000 markers. Imputation was performed using Beagle version 5.5[11] using a reference panel from The Ashkenazi Genome Consortium (TAGC, n = 128).[12] Variants with a dosage $R^2$, an imputation quality metric approximating the squared correlation between imputed dosages and true genotypes, below 0.8 were excluded. Because of the limited size of the reference panel, we did not apply an additional minor allele frequency threshold for rare variants. Scores were based on published weight files for Parkinson's disease and laryngeal dystonia (90 and 1804 markers respectively), with no additional LD clumping or p-value thresholding.[13–15] A polygenic risk score was calculated for each participant using PLINK2.0, with exclusion of missing genotypes.[16] Groups were compared using the Mann-Whitney U test for z-scored cumulative PRS values and Fisher’s exact test for participants at the extremes of the cumulative score distribution.

Phasing and quality control

VCF and BAM files from long-read sequencing were phased using WhatsHap,[17] using default parameters with indel phasing enabled. Variants that remained unphased by WhatsHap were subsequently phased with Beagle to maximise haplotype completeness using the TAGC reference panel.[12] WES and WGS were phased with Beagle 5.5[18] with 24 iterations, using TAGC as a reference.[12] Imputation with Beagle was only considered for the haplotype in *trans* to the c.907_909del, because this deletion was not present in the reference panel and therefore could not be directly imputed.

Presence of the variants was validated manually by visually inspecting the raw reads in the Integrative Genomic Viewer.[19] For quality control, we verified that c.646G>C occurred only in *trans* to c.907_909del. A cutoff of depth > 15 reads was used for verification of the phasing.

We lifted VCF coordinates between GRCh37/hg19 and GRCh38/hg38 using UCSC chain files, excluding non-unique or unmapped variants.

Short-read and long-read whole-genome sequencing were used to validate phasing and imputation results. Sequencing was prioritised in parent–offspring pairs or siblings with contrasting phenotypes, where differences in haplotype inheritance were expected to be most informative (Supplementary Figure 1).

#### Haplotype clustering and analysis

Analysis of the haplotypes was performed in R 2024.12.1. Alleles were coded as 0 for the reference alleles and 1 for the non-reference allele. Euclidean distance and complete linkage were used for hierarchical clustering. To aid cluster definition, we performed principal component analysis (PCA) using the prcomp function from the R stats package. Because PC1 explained most of the variance, groups were separated using data-driven breakpoints defined by local minima in the PC1 kernel density curve.

#### Population genetic analysis

Variant frequency was determined using gnomAD.[20] Analysis of linkage disequilibrium of variants in the minor haplotype was performed using LDlink[21] with the LDpair and LDmatrix tools in hg19, focusing on African, European and East Asian populations. Variants from Neanderthal and Denisovan DNA were examined using Ancient Genome Browser.[22] The age of each variant was calculated using the Human Genome Dating tool.[23]

#### *In silico* predicted expression

Analysis of Variant Expression quantitative trait loci (eQTL) was derived from the Adult Genotype Tissue Expression (GTEx) database using GTEx eQTL calculator[24] on ten representative variants (appendix table S5). Tissues examined were amygdala, anterior cingulate cortex, caudate, cerebellar hemisphere, cerebellum, cortex, frontal cortex, hippocampus, hypothalamus, nucleus accumbens, putamen, substantia nigra, cultured fibroblasts, EBV-transformed lymphocytes and whole blood.

For 12 participants who underwent long-read sequencing, we extracted from the *trans* haplotype 1,048,576-bp sequences centred on the coordinates of c.907_909del. Regulatory activity was predicted with the AlphaGenome DNA model.[25] For each sample, tissue-specific RNA-seq activity was estimated for cerebellum, caudate nucleus, putamen and whole blood using UBERON terms,[26] *TOR1A*/*TOR1B* expression was quantified across GENCODE v46 gene models.

#### Blood DNA Methylation analysis

5mC modifications on CpG sites from ONT were assessed using modkit (Oxford Nanopore Technologies) and compared in the 30 kb regions surrounding the c.907_909del variant using Student's t-test. In addition, visual assessment of 5mC modification on CpG sites +/- 1,000 bp around the 3'UTR of each gene was performed using IGV desktop, methylated/unmethylated promoters were compared using Fisher's exact test.

#### PBMC/LCL Processing and qPCR Expression Analysis

Peripheral blood (10 mL per donor) was collected into EDTA tubes for whole-blood analyses and into heparin tubes for lymphoblastoid cell line (LCL) generation. Samples were processed within 1h of collection and maintained at 2–8 °C during transport. Peripheral blood mononuclear cells (PBMCs) were isolated by density-gradient centrifugation using Lymphoprep (STEMCELL Technologies) according to the manufacturer's instructions.

For LCLs, PBMCs were transformed with Epstein–Barr virus (EBV) in the presence of cyclosporin A. Cultures were maintained at 37 °C, 5% $CO_2$ in Roswell Park Memorial Institute (RPMI) 1640 supplemented with 20% FBS, 1% L-glutamine, and 1% penicillin/streptomycin (Biological Industries Beit Haemek, Israel). LCLs were expanded for 3-4 weeks until stable. For expression analyses, ~$2\times10^6$ cells were seeded and cultured for 48 h prior to RNA extraction.

Total RNA was extracted from PBMCs/LCLs using TRIzol (Thermo Fisher) following the manufacturer's protocol. Quantity and purity were assessed by NanoDrop (A260/280 and A260/230 ratios). cDNA was synthesised from 1,000 ng RNA using the qScript cDNA Synthesis Kit (Quantabio) according to the manufacturer's instructions.

qPCR was performed using PerfeCTa SYBR Green FastMix ROX (Quantabio) in 10 µL reactions in the QuantaStudio 5 Real-Time PCR system (Thermo Fisher). Primers targeted the *TOR1A* c.907_909del allele, the *TOR1A* wild-type allele, and *TOR1B* (see appendix table S6 for sequences). The specificity of the mutant allele-specific primer was validated using samples from mutation carriers and a non-carrier control. *RPLP0* and *GUSB* served as reference genes after stability verification under our conditions. Each target was run with three technical replicates. For LCL experiments, one representative participant-derived cell line from each haplotype group was studied, with three repeated culture-based measurements per line. No-template controls were included in each run. Melt-curve profiles were single-peaked, supporting amplicon specificity. Reactions with failed amplification or non-specific melt-curve profiles were excluded according to predefined quality-control criteria.

Cq values were averaged across technical replicates after exclusion of outliers due to failed amplification or melt-curve artefacts (pre-specified rules). Relative expression was calculated using the $2^{-\Delta\Delta Ct}$ method. Statistical analysis was performed using Student's t-test for blood samples comparing Haplotype A and Haplotype B (as only one sample was available in the mixed haplotype group), and the Jonckheere–Terpstra test for biological replicates from LCLs to assess an a priori ordered trend across haplotype groups.

### SUPPLEMENTARY TABLES

Table S1: Clinical severity scoring system used in the study

| Score | Group in main analysis | Criteria |
|---|---|---|
| 0 | minDys | Asymptomatic individuals older than 26 years as the onset of DYT-*TOR1A* is rare in older individuals.[27,28] |
| 1 | | Individuals older than 26 years with minimal signs of dystonia such as mild action-specific non-disabling dystonia of a single upper-limb (e.g. writer's cramp) or low amplitude tremor.[28,29] |
| 2 | Excluded | Participants older than 16 years with upper limb, trunk, neck or face dystonia induced by multiple actions or observed at rest without lower limb involvement, a clinical picture associated with less predictable life-long prognosis.[28,29] |
| 3 | svrDys | Generalised dystonia involving the lower limbs with impaired but independent ambulation. |
| 4 | | Generalised dystonia involving the lower limbs with loss of independent ambulation. |

Table S2: Primers used for PCR amplification and Sanger sequencing

| Variant(s) | Forward (5'->3') | Reverse (3'->5') |
|---|---|---|
| rs753167620 (c.907_909del) | CCTGGAATACAAACACCTA | GGCTGCCAATCATGACTGTC |
| rs1801968 (c.646G>C) | GACAACGCGTGTTCAATGTC | CCTCTGAGCTCCAGGAGAAA |
| rs3842225 | TAGGAGGTTTCGGGGATTCT | CTGAAACAGCCTCTCCAAGG |
| rs13297609 | ACCTGGAGTTCATCAGCAAAG | GGAGTGGAAAGCAGAGGGAA |
| rs13294595 | GTCACTTACCTACCCTCCCA | GGTGCCTGTTAACATCTGCC |
| rs10988519 | CCTTCGCAAACAGTCTCCAG | CAAATGTGGGCTTTCTGGGC |
| rs34814665, rs112575920 | TCCCACGGCCAACTTCTATT | CCTCCCTATCTCAGTTGCGA |
| rs13300897 | GTAGAGACGCGGGTAGATGT | AACAGGGCTTTGTACCGAAC |
| rs1590705, rs11788537 | GCATTAGAAACTGGCTGGGC | AGTTGGAGATGTCAGCACGA |

Table S3: Distribution of 5-variant genotype combinations in the source retrospective cohort

| Five-variant genotype combination | rs2296793 | rs1801968 | rs3842225 | rs2287367 | rs1043178 | Ashkenazi-Jewish | | Non-Jewish | |
|---|---|---|---|---|---|---|---|---|---|
| | | | | | | ΔGAG | non-ΔGAG | ΔGAG | non-ΔGAG |
| Haplotype A associated combination | G/G | G/G | G/G | G/G | T/C | 53 | 12 | 16 | 11 |
| | G/G | G/G | G/G | G/G | T/T | 46 | 8 | 10 | 1 |
| c.646G>C associated combination | G/G | C/G | G/G | G/G | T/C | 16 | 6 | 0 | 2 |
| Haplotype B associated combination | A/G | G/G | G/- | T/G | T/C | 32 | 10 | 4 | 6 |
| Other genotype combinations | - | - | - | - | - | 1 | 42 | 60 | 27 |
| Total | - | - | - | - | - | 148 | 78 | 90 | 47 |
| Matching a combination identified in the original cohort, % | - | - | - | - | - | 99.3% | 46.2% | 33.3% | 42.6% |

Genotype data for five variants were available in the independent retrospective cohort. Five-locus genotype combinations were compared with the phased, extended haplotypes characterised by whole-exome, whole-genome, and long-read sequencing in the original cohort. Variants rs2296793, rs3842225, and rs2287367 are located within a relatively short segment of haplotype B. This marker combination was also observed in individuals with a mixed haplotype. Variant rs1801968 corresponds to c.646G>C, and rs1043178 was phased *in cis* with c.907_909del (ΔGAG).

Among Ashkenazi Jewish c.907_909del carriers, 147 of 148 (99.3%) had one of the four genotype combinations identified in the original cohort. The remaining carrier had missing genotype data and was excluded from the analysis. In contrast, only 30 of 90 (33.3%) non-Jewish c.907_909del carriers had one of these combinations, indicating substantially greater diversity in the genetic backgrounds carrying c.907_909del. Because the analysis evaluated the protective effect of the extended haplotype as a linked set of variants rather than the effects of individual variants, the original haplotypes could not be reliably inferred from these five markers in the non-Jewish population. Non-Jewish carriers were therefore excluded from the study.

Table S4: Genes used in our panels for Dystonia and Parkinson's disease

| Panel | Genes |
|---|---|
| Dystonia (38 genes) | *ANO3, AOPEP, ATP13A2, ATP1A3, CP, DCTN1, DNAJC6, EIF2AK2, FBXO7, GBA1, GCH1, GLB1, GNAL, HPCA, KCTD17, KMT2B, LRRK2, PARK7, PINK1, PLA2G6, PRKN, PRKRA, PTS, QDPR, RAB32, SGCE, SLC30A10, SLC6A3, SNCA, SPR, SYNJ1, TAF1, TH, THAP1, TOR1A, VPS13C, VPS16, VPS35* |
| Parkinson's disease (101 genes) | *ANO3, APP, CSF1R, GBA1, GCH1, GIGYF2, GNAL, EIF4G1, C9orf72, C19orf12, GRN, DNAJB2, DNAJC5, DNAJC6, DNAJC12, DNAJC13, ATN1, ATP1A3, ATP6AP2, FTL, FUS, ATP7B, ATP13A2, ATXN1, ATXN2, ATXN3, OPA3, PINK1, PLA2G6, TOR1A, TMEM230, SLC20A2, SLC30A10, SLC39A14, SLC41A1, PPP2R2B, PRKN, PRKRA, SYNJ1, TAF1, TARDBP, TBP, RAB39B, CHCHD2, CHCHD10, CLN3, COASY, CP, CYP27A1, DCAF17, DCTN1, HTRA2, HTT, LRP10, LRRK2, LYST, FBXO7, IPPK, JAM2, JPH3, KIF5A, MAPT, NPC1, NPC2, NR4A2, NUS1, PDE8B, PDE10A, PDGFB, PDGFRB, PANK2, PARK7, MYORG, MT-TY, POLG, POLG2, PSEN1, PSEN2, PTRHD1, PTS, SGCE, SLC6A3, SMPD1, SNCA, SNCAIP, SNCB, SPG11, SPR, TENM4, TH, THAP1, TUBB4A, TWNK, UCHL1, VAC14, VPS13A, VPS13C, VPS35, WDR45, XPR1, ZFYVE26* |

Table S5: Representative variants from the studied haplotypes

| rsID | Pos (hg38) | Ref | Alt | Gene | AF in AJ | AF in European | AF in African | Altai Neanderthal | Ldpair with rs67061074 (p-value<0.0001) | Ldpair with rs67061074 in Europeans (p-value<0.0001) | Ldpair with rs67061074 in East Asians (p-value<0.0001) | Ldpair with rs67061074 in Africans (p-value<0.0001) | Age in generations according to human.genome.dating | Age in years |
|---|---|---|---|---|---|---|---|---|---|---|---|---|---|---|
| rs67061074 | 9:129833880 | C | T | C9orf78 | 0.2726 | 0.2411 | 0.2008 | Yes | | | | | 51,355.40 | 1,283,885 |
| rs7032945 | 9:129829093 | T | G | C9orf78 | 0.2748 | 0.2430 | 0.2182 | Yes | Yes | Yes | Yes | Yes | 43,241.70 | 1,081,043 |
| rs35168956 | 9:129828558 | G | A | C9orf78 | 0.2833 | 0.2401 | 0.2198 | No | Yes | Yes | Yes | Yes | 45,491.30 | 1,137,283 |
| rs13300897 | 9:129824189 | C | T | Intergenic | 0.2061 | 0.2247 | 0.04792 | No | Yes | Yes | Yes | No | 7,129.60 | 178,240 |
| rs13297609 | 9:129818407 | G | C | *TOR1A* | 0.1985 | 0.1869 | 0.04155 | No | Yes | Yes | Yes | No | 13,170.50 | 329,263 |
| rs13283469 | 9:129819735 | C | T | *TOR1A* | 0.1981 | 0.2190 | 0.04925 | No | Yes | Yes | Yes | No | 7,225.30 | 180,633 |
| rs13283293 | 9:129812198 | G | A | Intergenic | 0.2869 | 0.2518 | 0.2408 | Yes | Yes | Yes | Yes | Yes | 43,562.80 | 1,089,070 |
| rs11789668 | 9:129807471 | C | G | *TOR1B* | 0.2078 | 0.2248 | 0.04887 | No | Yes | Yes | Yes | No | 6,574.30 | 164,358 |
| rs11788134 | 9:129804914 | G | A | *TOR1B* | 0.1920 | 0.1873 | 0.04164 | No | Yes | Yes | Yes | No | 8,243.00 | 206,075 |
| rs7025334 | 9:129794412 | T | C | Intergenic | 0.1781 | 0.1895 | 0.05528 | No | Yes | Yes | Yes | No | 19,852.80 | 496,320 |

Representative variants were chosen to span the haplotype interval at approximate spacing, prioritising loci well covered in exome data and variants within or near regional genes. Age was calculated assuming 25 years per generation. Abbreviations: rsID: Reference SNP cluster ID. Pos: position in hg38. Ref: reference base. Alt: Alternative base in the minor haplotype. AF: Allele frequency. AJ: Ashkenazi Jewish. LDpair: linkage disequilibrium of both minor alleles in a given population.

#### Table S6: Primers used for real-time PCR

| Gene | Forward (5'->3') | Reverse (3'->5') |
|---|---|---|
| *TOR1A*$_{wt}$ | CACCTAAAAATGTGTATCCGAGT | TTTGGGGAAAAATGTCATCTCCTC |
| *TOR1A*$_{mut}$ | CACCTAAAAATGTGTATCCGAGT | TTTGGGGAAAAATGTCATCTCAGC |
| *TOR1B* | ACGGAGTGTCTTACCGCAAAGC | ACAGGTTCCAGGTCCTTCAGCT |
| *RPLP0* | GAAACTCTGCATTCTCGCTTCC | GACTCGTTTGTACCCGTTGATG |
| *GUSB* | TCCAAATGAGCTCTCCAACC | AAACGATTGCAGGGTTTCAC |

#### Table S7: Logistic regression analysis for sex, c.646G>C and Haplotype B

| | Odds ratio | Lower 95% | Upper 95% | P-value |
|---|---|---|---|---|
| **Model A** | | | | |
| Male sex | 1.67 | 0.93 | 3.00 | .094 |
| c.646G>C | 0.08 | 0.01 | 0.29 | **.001** |
| Haplotype B | 0.47 | 0.23 | 0.92 | **.030** |
| **Model B** | | | | |
| Male sex | 1.66 | 0.94 | 2.98 | .083 |
| c.646G>C or Haplotype B | 0.31 | 0.16 | 0.58 | **<.001** |
| **Model C** | | | | |
| c.646G>C or Haplotype B | 0.32 | 0.17 | 0.60 | **<.001** |

A) The dependent variable is defined as any severity level above 1. Male sex is associated with higher risk, even if not significantly (p=0.094). In contrast, both c.646G>C and Haplotype B are significantly associated with reduced penetrance (p=0.001 and p=0.030 respectively). B) When combined, having either c.646G>C or Haplotype B (mutually exclusive when in *trans* to c.907_909del) has a stronger association with penetrance (p<0.001) while sex is still not significant (p=0.083). C) Same as (B), but when the only independent variable is having a protective haplotype. OR: odds ratio.

Table S8: Average normalised effect size (NES) of representative variants in Haplotype B on expression of *TOR1A*, *TOR1B* and the difference between them, analysed by GTEx eQTL calculator.

| Tissue | *TOR1A* | *TOR1B* | *TOR1B*-*TOR1A* difference |
|---|---|---|---|
| Brain_Cerebellar_Hemisphere | -0.733** | 0.568** | 1.301 |
| Brain_Cerebellum | -0.81** | 0.385** | 1.195 |
| Brain_Caudate_basal_ganglia | -0.196** | 0.543** | 0.739 |
| Brain_Putamen_basal_ganglia | -0.173 | 0.544** | 0.717 |
| Brain_Amygdala | -0.184** | 0.42* | 0.604 |
| Brain_Nucleus_accumbens_basal_ganglia | -0.1244 | 0.466** | 0.5904 |
| Brain_Anterior_cingulate_cortex_BA24 | -0.126 | 0.402* | 0.528 |
| Brain_Frontal_Cortex_BA9 | -0.133 | 0.373* | 0.506 |
| Brain_Cortex | -0.181** | 0.316* | 0.497 |
| Brain_Hypothalamus | -0.161** | 0.332** | 0.493 |
| Brain_Hippocampus | -0.123 | 0.341* | 0.464 |
| Brain_Substantia_nigra | -0.175** | 0.1708** | 0.3458 |
| Brain_Spinal_cord_cervical_c-1 | -0.1176 | 0.186** | 0.3036 |
| Cells_EBV-transformed_lymphocytes | 0.036956 | 0.322 | 0.285044 |
| Cells_Cultured_fibroblasts | 0.0058 | -0.0976* | -0.1034 |
| Whole_Blood | -0.26** | -0.427** | -0.167 |

* adjusted p-value<0.05 in most variants (> 5/10) ** adjusted p<0.05 in all variants (10/10)

Table S9: Mixed haplotypes in two patients carry only a subset of haplotype B alleles across representative variants.

| rsID | ID | Hap. A | Hap. B | Patient 11-II-1 (Mixed Hap.) | Patient 3-II-2 (Mixed Hap.) | *TOR1A* caudate - NES | *TOR1B* caudate - NES | *TOR1A* cerebellum - NES | *TOR1B* cerebellum - NES |
|---|---|---|---|---|---|---|---|---|---|
| rs67061074 | 9:129833880-C>T | C | **T** | **T** | C | -0.21 | 0.65 | -0.81 | 0.39 |
| rs7032945 | 9:129829093-T>G | T | **G** | **G** | T | -0.21 | 0.65 | -0.82 | 0.39 |
| rs35168956 | 9:129828558-G>A | G | **A** | **A** | G | -0.21 | 0.65 | -0.82 | 0.39 |
| rs13300897 | 9:129824189-C>T | C | **T** | **T** | **T** | -0.19 | 0.53 | -0.79 | 0.39 |
| rs13297609 | 9:129818407-G>C | G | **C** | G | **C** | -0.19 | 0.43 | -0.82 | 0.37 |
| rs13283469 | 9:129819735-C>T | C | **T** | **T** | **T** | -0.19 | 0.52 | -0.81 | 0.39 |
| rs13283293 | 9:129812198-G>A | G | **A** | **A** | **A** | -0.21 | 0.63 | -0.83 | 0.4 |
| rs11789668 | 9:129807471-C>G | C | **G** | **G** | **G** | -0.18 | 0.52 | -0.8 | 0.39 |
| rs11788134 | 9:129804914-G>A | G | **A** | G | **A** | -0.18 | 0.42 | -0.81 | 0.37 |
| rs7025334 | 9:129794412-T>C | T | **C** | T | **C** | -0.19 | 0.43 | -0.79 | 0.37 |

Haplotypes (Hap.) A and B are shown as reference allelic patterns for the 10 representative variants. The alleles present in the two mixed haplotypes are shown for comparison. Mixed haplotypes 1 and 2 each carry only a subset of haplotype B alleles, such that neither reconstructs the complete haplotype B background. In patient 3-II-2 variants were validated using Sanger sequencing with a clear recombination between Haplotype A and Haplotype B between rs35168956 and rs13300897. In contrast, in patient 11-II-1 the variants, confirmed with whole genome sequencing with high reading depth, have shown a patchy mixture between Haplotype A and B. This may account for the fact that patient 11-II-1 inherited this trans haplotype from a non-Jewish parent, thus this may be a unique haplotype that was not represented in the reference cohorts. Corresponding *TOR1A* and *TOR1B* normalised effect sizes (NES) in caudate and cerebellum are shown. All haplotypes presented are *in trans* to c.907_909del.

Table S10: Methylation analysis in whole blood of carriers of Haplotype A and Haplotype B

| Haplotype | No. of samples | Mean sample methylation ± SD | Samples with overall *TOR1A* promoter methylation (n/N) | Samples with overall *TOR1B* promoter methylation (n/N) |
|---|---|---|---|---|
| A | 10 | 0.685 ± 0.025 | 0/10 | 0/10 |
| B | 2 | 0.682 ± 0.006 | 0/2 | 0/2 |
| P-value (A vs. B) | | **0.755** | **1.000** | |

Whole blood DNA methylation status by haplotype among participants who underwent Nanopore long-read sequencing, including quantitative promoter methylation measures and overall promoter methylation status based on visual inspection in IGV desktop (see methods). Mean methylation was compared between haplotypes using Student's t-test, and categorical promoter methylation status was compared using Fisher's exact test. Abbreviations: SD: Standard deviation.

Table S11: Demographic, clinical, and genetic characteristics of participants with lymphoblastoid cell lines (LCLs).

| Participant | *Trans* Protective haplotype B status | Age range (years) | Sex | Clinical Score | Age at symptom onset (years) | *TOR1A* c.646G>C status |
|---|---|---|---|---|---|---|
| 16-III-1 | Full | 35-39 | Male | 1 | N/A | No |
| 11-II-1 | Mixed | 40-44 | Female | 4 | 7 | No |
| 16-III-2 | Absent | 30-34 | Male | 3 | 12 | No |

### SUPPLEMENTARY FIGURES

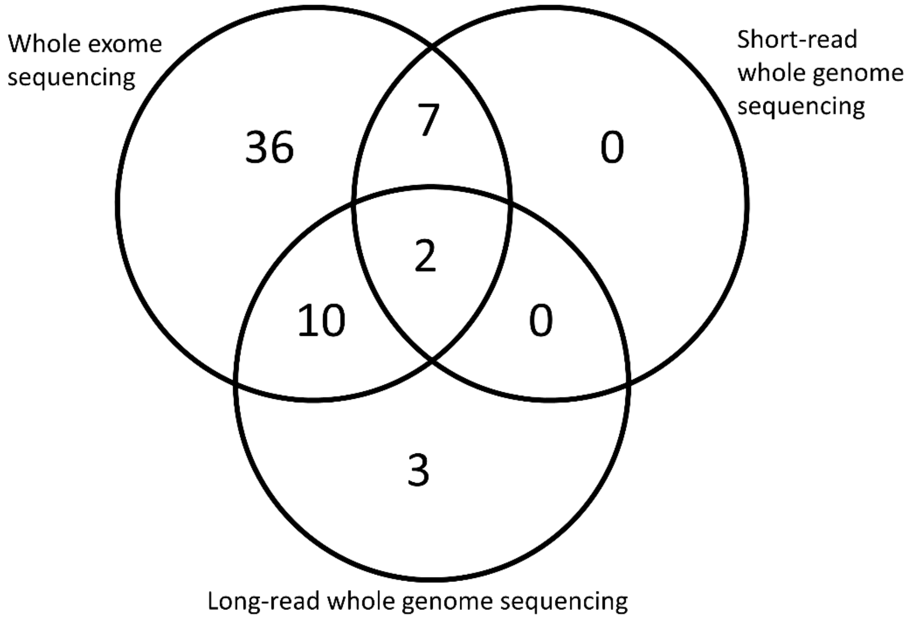


Figure S1**. A Venn diagram of different sequencing techniques used.** A Venn diagram demonstrating the number of *TOR1A* c.907_909del carriers performing Whole exome sequencing (total n = 54), short-read whole genome sequencing (total n = 9) and long-read whole genome sequencing (total n = 15).

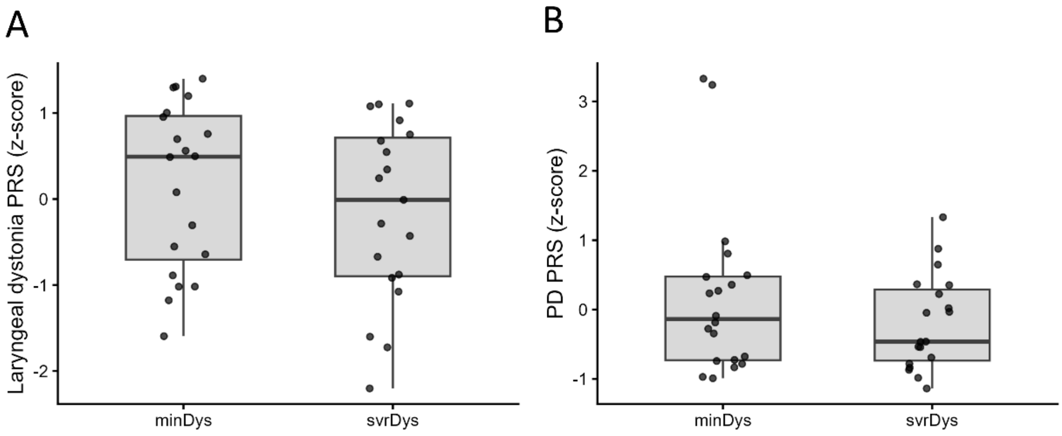


Figure S2**. Polygenic risk scores (PRS) for laryngeal dystonia and Parkinson's disease do not explain penetrance.** A) Laryngeal focal dystonia PRS are similar between minDys (median 0.493) and svrDys (median −0.009; p=0.332). A categorical comparison between the bottom vs. top 20% of PRS showed no difference (P = 1.000). The central black line in the boxplot marks the median value; the box spans the interquartile range (IQR; 25th–75th percentiles). B) Parkinson's disease PRS are similar between minDys (median −0.138) and svrDys (median −0.462; p=0.491). A categorical comparison of disease severity between the bottom vs. top 20% of PRS also showed no difference (p=1.000).

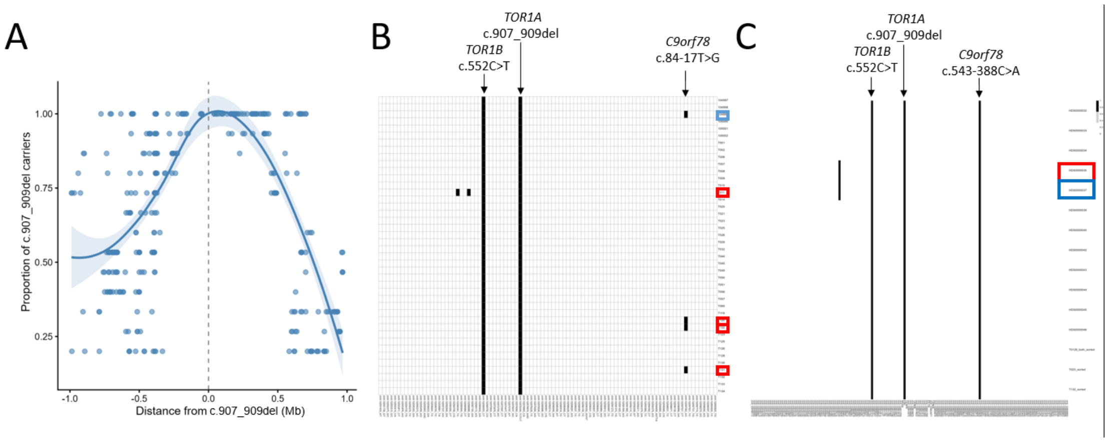


Figure S3: **Lack of modifier variants in *cis* of *TOR1A* c.907_909del**. A) Visualisation of rare variants (Allele frequency, AF < 0.05 in TAGC) in *cis* to the c.907_909del that are present in more than two participants, demonstrating the outlines of the cis allele. B) Among minDys and svrDys patients who underwent WES (N = 42), the analysis of the 50kb in cis to the c.907_909del, a region that encompasses *TOR1B*, *TOR1A*, *C9orf78* and part of *USP20*, found only two additional variants in more than one participant: c.552C>T (p.Pro184Pro) in *TOR1B* (rs1043178, chr9:129807274) present in all participants, and c.84-17T>G in *C9orf78* (rs41278736, chr9:129834783) that was detected in four participants and showed no association with dystonia (p=0.178). The x-axis shows rare variants (AF < 0.05 in TAGC) in cis to TOR1A c.907_909del that were detected in more than two participants, and the y-axis shows individual participants. Filled black rectangles correspond to the presence of the variant. C) Analysis of the same cis region among 15 participants using ONT showed another shared variant in C9orf78 (rs41278734, chr9:129829929, c.543-388C>A) and a single intragenic variant (rs530319119, chr9:129798334) present in only two individuals (p=0.377). Together, these results support the absence of cis-acting modifiers linked to the c.907_909del variant in the proximal region. Abbreviations: minDys: minimal dystonia (blue boxes). svrDys: severe dystonia (red boxes).

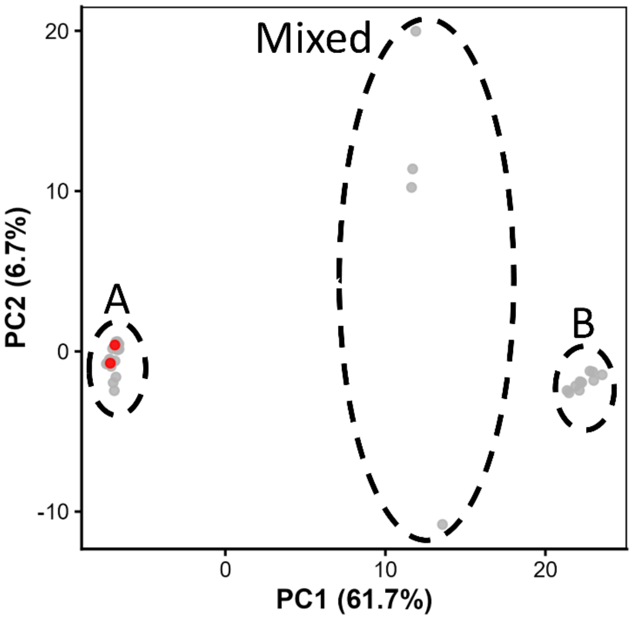


Figure S4: **Principal component analysis of sequenced participants.** Principal component analysis (PCA) was performed using variants within a 50-kb region *in trans* to *TOR1A* c.907_909del among sequenced participants included in the analysis dataset (n = 53). The analysis demonstrated clear separation along PC1 into three major subgroups, accounting for 61.7% of the observed genetic variation. Participants carrying *TOR1A* c.646G>C are shown in red and cluster within Haplotype A. A indicates Haplotype A; B indicates Haplotype B; and Mixed indicates individuals with a combination of haplotypes A and B. Note: One additional participant was sequenced after completion of the PCA analysis and was therefore not included in this figure.

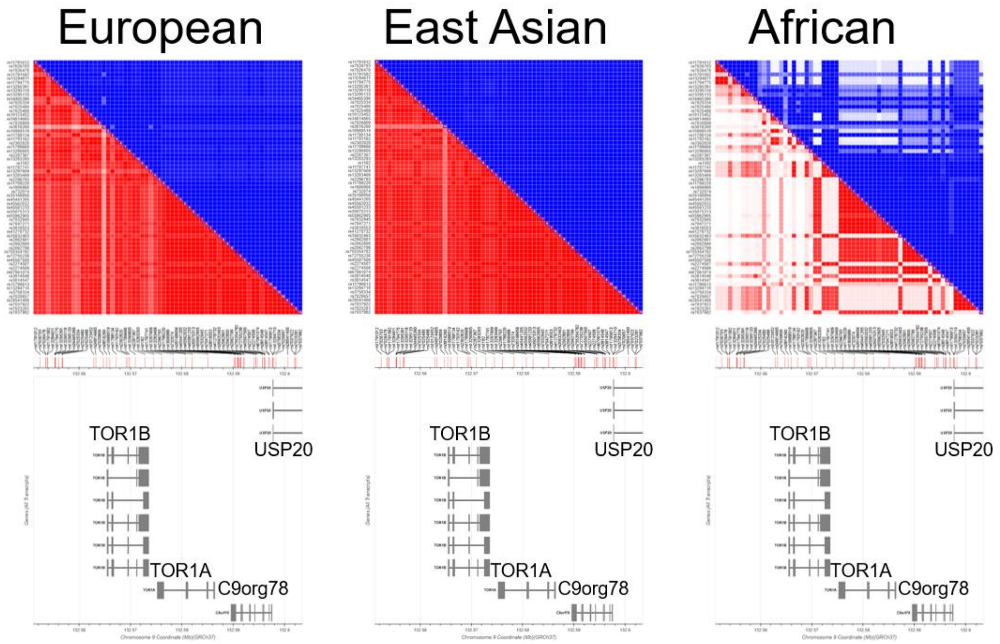


Figure S5: **LD plot of different populations reveals two distinct chronological origins of Haplotype B**. A correlation plot demonstrating $R^2$ (Squared Correlation Coefficient, red) and D' (Normalised Coefficient, blue) for variants of Haplotype B. While almost all variants show a high degree of correlation in European and East Asian populations, in African populations only certain variants, mostly around *TOR1A*, *C9orf78* and a small part of *TOR1B* are strongly correlated. LD: linkage disequilibrium. The figure was produced with LDlink (see methods).

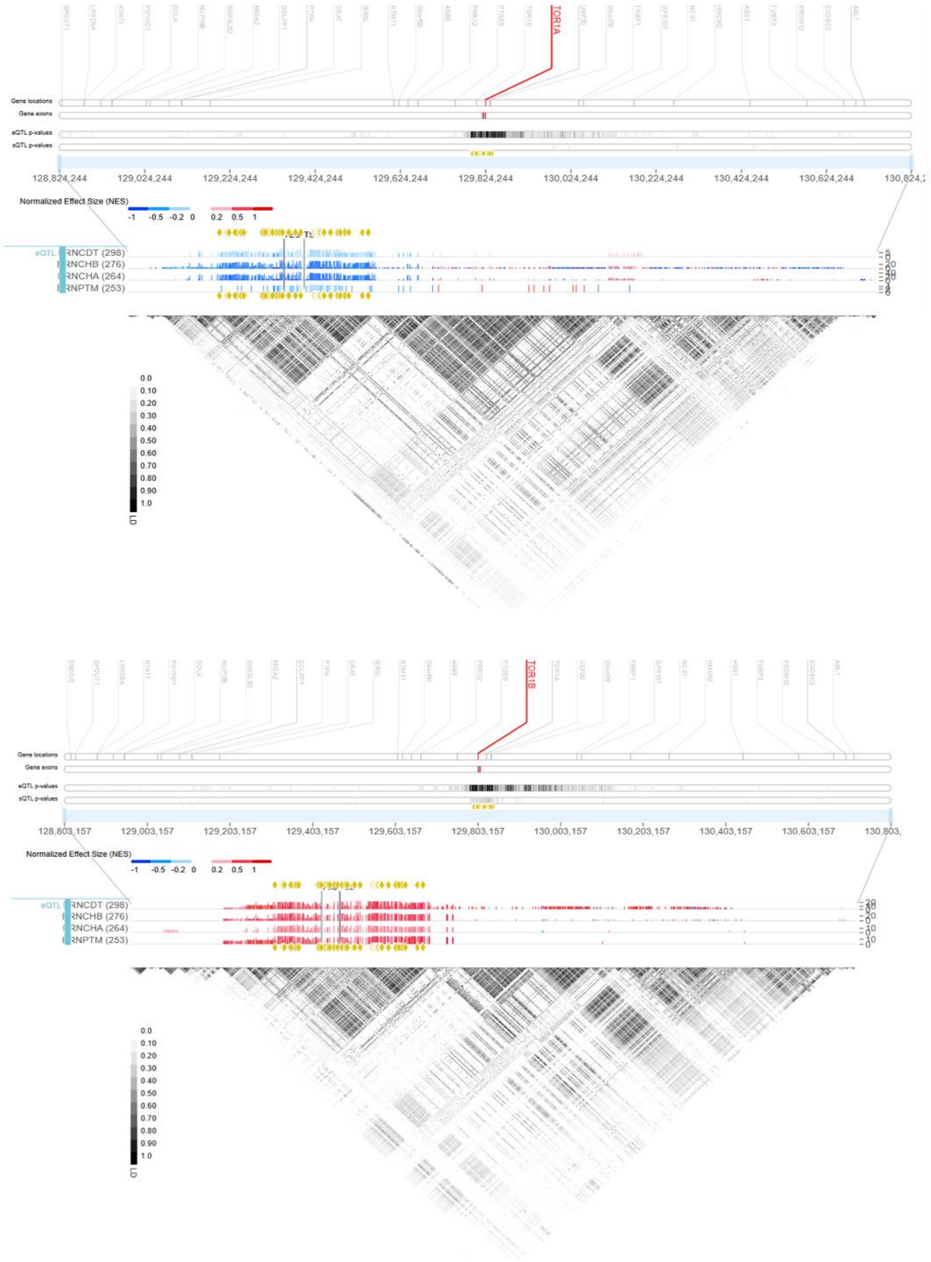


Figure S6. **eQTL mapping across a 200-kb region**. eQTL mapping of variants comprising Haplotype B. Across a 200-kb region, variants comprising Haplotype B (yellow rectangles) are located within a single linkage-disequilibrium (LD) block, with darker shading indicating stronger LD. Variants within this block are the main determinants of reduced TOR1A expression (upper panel) and increased TOR1B expression (lower panel) in relevant brain tissues. Blue indicates a negative normalised effect size, corresponding to stronger downregulation, whereas red indicates a positive normalised effect size, corresponding to stronger upregulation. BRNCDT, caudate nucleus; BRNCHB, cerebellar hemisphere; BRNCHA, cerebellum; BRNPTM, putamen. The figure was generated using the GTEx Locus Browser.

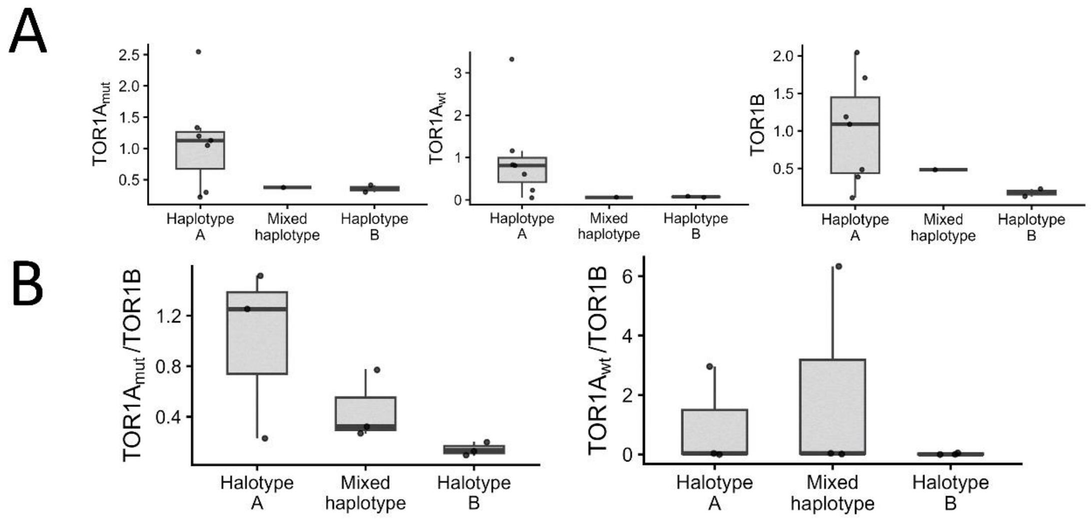


Figure S7: **Association between Haplotype B and the expression of the mutant TOR1A allele in Blood and patients-derived lymphoblastoid cell lines.** A) In peripheral blood, relative expression (fold change vs, mean Haplotype A) shows a significant decrease in both the mutant TOR1A allele ($TOR1A_{mut}$, p=0.042) and TOR1B (p=0.022) among carriers of Haplotype B (n=2) compared to carriers of Haplotype A (n=7), with the mixed haplotype having intermediate values. $TOR1A_{wt}$ likewise showed a nonsignificant trend towards lower expression (p=0.065). B) In lymphoblastoid cell lines from three representative participants, the $TOR1A_{mut}$ /TOR1B ratio showed a significant dose-dependent decrease with Haplotype B (p=0.011), driven primarily by attenuation of the mutant allele (p=0.005) with no significant change in TOR1B (p=0.741). By contrast, the $TOR1A_{wt}$/TOR1B ratio did not show a significant change across haplotypes (p=0.334). The central black line in the boxplot marks the median value; the box spans the interquartile range (IQR; 25th–75th percentiles).
